# Identification of multiple large deletions in ORF7a resulting in in-frame gene fusions in clinical SARS-CoV-2 isolates

**DOI:** 10.1101/2020.06.08.20125856

**Authors:** Amin Addetia, Hong Xie, Pavitra Roychoudhury, Lasata Shrestha, Michelle Loprieno, Meei-Li Huang, Keith R. Jerome, Alexander L. Greninger

## Abstract

Peculiar among human RNA viruses, coronaviruses have large genomes containing accessory genes that are not required for replication. Numerous mutations within the SARS-CoV-2 genome have been described but few deletions in the accessory genes of SARS-CoV-2 have been reported. Here, we report two large deletions in ORF7a, both of which produce new open reading frames (ORFs) through the fusion of the N-terminus of ORF7a and a downstream ORF.

---

Peculiar among human RNA viruses, coronaviruses have large genomes containing accessory genes that are not required for replication (1). Numerous mutations within the SARS-CoV-2 genome have been described (2) but few deletions in the accessory genes of SARS-CoV-2 have been reported (3, 4). Here, we report two large deletions in ORF7a, both of which produce new open reading frames (ORFs) through the fusion of the N-terminus of ORF7a and a downstream ORF.

We recovered genomes from the SARS-CoV-2 isolates from two separate patients as part of an ongoing University of Washington IRB-approved genomic surveillance project (5, 6). Sequencing libraries were generated from double-stranded cDNA (7) using Swift Biosciences’ SARS-CoV-2 Normalase Amplicon Sequencing kit or the Nextera DNA Flex Pre-Enrichment kit (Illumina) followed by enrichment using a SARS-CoV-2 xGen enrichment panel (NC_045512; IDT). Sequence reads were adapter- and quality-trimmed using Trimommatic v0.38 (8), aligned to the SARS-CoV-2 reference genome (NC_045512.2) using BBMap (https://sourceforge.net/projects/bbmap/), trimmed of synthetic PCR primers using Primerclip (https://github.com/swiftbiosciences/primerclip) if appropriate, and visualized in Geneious v11.1.4 (9). Sequencing reads and consensus genomes are available under NCBI BioProject PRJNA610428.

In WA-UW-4570, we identified a 392-nucleotide deletion originating at nt 27,494 in ORF7a. This deletion results in the loss of the entirety of ORF7b and creates a new ORF through the fusion of the N-terminus of ORF7a with ORF8 (Figure 1a). Notably, this is the largest deletion recorded to date in SARS-CoV-2. In WA-UW-5812, we identified a 227 nt deletion also resulting a new ORF through the fusion of the N-terminus of ORF7a with ORF7b (Figure 1b). Both deletions were confirmed by RT-PCR (Figure 1c-d) and Sanger sequencing. In contrast to a previously sequenced SARS-CoV-2 isolate with a deletion in ORF7a (3), our isolates retained the presumptive signal peptide and part of the β sheet of the Ig superfamily fold (10) of ORF7a (Figure 1e-f) and lost the protein’s transmembrane and cytoplasmic domains (PDB 6W37) (10).

**Figure 1.**
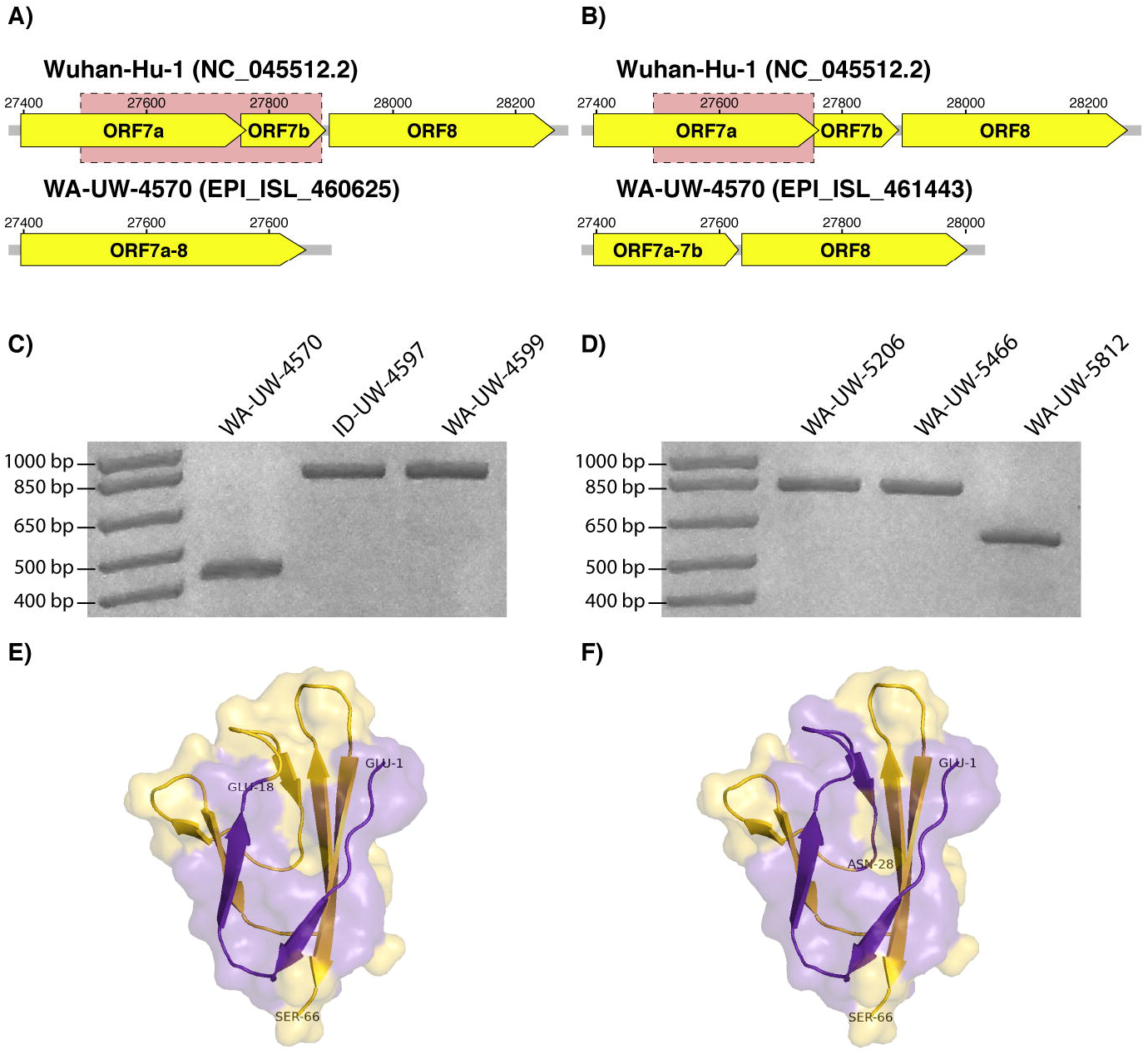
A 392-nucleotide deletion starting at nt 29,424 was identified in **a)** WA-UW-4570 and resulted in the fusion of ORF7a and ORF8. A 227-nucleotide deletion beginning at nt 27,524 was identified in **b)** WA-UW-5812 and resulted in the fusion of ORF7a and ORF7b. The deletions in **c)** WA-UW-4570 and **d)** WA-UW-5812 were confirmed by RT-PCR. An 826-bp product is expected for strains with an intact ORF7a. 434-bp and 599-bp amplicons were obtained for WA-UW-4570 and WA-UW-5812, respectively. The deletions in **e)** WA-UW-4570 and **f)** WA-UW-5812 severely truncate the β sheet and result in the loss of the transmembrane and cytoplasmic domains of ORF7a (PDB 6W37) (10). Residues retained are highlighted in purple, while those lost are highlighted in gold.

We next assessed the genetic relatedness of the two strains. WA-UW-4570 and WA-UW-5812 differed by 12 single nucleotide variants and a 3-bp deletion in the spike protein of WA-UW-4570 and belonged to the SARS-CoV-2 phylogenetic lineages A.1 and B.1 respectively (https://github.com/hCoV-2019/pangolin) (11). This suggests that the ORF7a deletions arose independently in the two strains and that deletions are likely not restricted to specific SARS-CoV-2 lineages. Both samples were recovered from the only SARS-CoV-2-positive nasopharyngeal swab available from each patient. The ORF1ab qRT-PCR cycle thresholds on the Hologic Panther Fusion for these isolates were 27.6 (WA-UW-4570) and 28.5 (WA-UW-5812). WA-UW-4570 was collected from an individual prophylactically taking hydroxychloroquine and mycophenolate mofetil while the WA-UW-5812 individual was not taking any medications for SARS-CoV-2 infection.

ORF7a of SARS-CoV-2, which is non-essential for growth *in vitro* (12), interacts with the ribosomal transport proteins HEATDR3 and MDN1 (13), and has been demonstrated to inhibit cellular translation in SARS-CoV (13). Based on the size of the deletions recovered and structure of ORF7a, we hypothesize that most biochemical functions of ORF7a would be inactivated by these deletions. Interestingly, ORF6 of SARS-CoV-2 interacts with the mRNA export proteins NUP98 and RAE1, and may additionally inhibit cellular translation (13). This redundancy may partially explain the ORF7a deletions observed in our isolates. We predict global sequencing projects may yield additional clinical SARS-CoV-2 isolates with deletions in ORF6 or ORF7a, but not both.

## Data Availability

Sequencing reads and consensus genomes are available under NCBI BioProject PRJNA610428.

